# SARS-CoV-2 IgG detection in human oral fluids

**DOI:** 10.1101/2021.07.07.21260121

**Authors:** Katja Hoschler, Samreen Ijaz, Nick Andrews, Sammy Ho, Steve Dicks, Keerthana Jegatheesan, John Poh, Lenesha Warrener, Thivya Kankeyan, Frances Baawuah, Joanne Beckmann, Ifeanichukwu O Okike, Shazaad Ahmad, Joanna Garstang, Andrew J Brent, Bernadette Brent, Felicity Aiano, Kevin E Brown, Mary E Ramsay, David Brown, John V Parry, Shamez N Ladhani, Maria Zambon

**Author notes:** **Correspondence:** Dr Katja Hoschler, Virus Reference Department, Public Health England, 61 Colindale Avenue, London NW9 5EQ, UK. **CONTRIBUTION** KH, SI, SH, TK, KJ and JP developed the assays and performed the laboratory analyses. NA and JVP performed the data analysis. SNL, FB, JB, IOO, SA, JG, AJB, BB, KEB, and MER devised, set up and recruited the sKIDs cohort and initiated and completed all the clinical sampling associated with sample acquisition. SL, JP, DB, SI, MZ and KH led on data interpretation and writing of the manuscript. All authors reviewed the data, discussed the results and commented on the manuscript. **FUNDING** This surveillance was funded by the Department of Health and Social Care (DHSC).

## Abstract

Seroepidemiological studies to monitor antibody kinetics are important for assessing the extent and spread of SARS-CoV-2 in a population. Non-invasive sampling methods are advantageous to reduce the need for venepuncture, which may be a barrier to investigations particularly in paediatric populations. Oral Fluids are obtained by gingiva-crevicular sampling from children and adults and are very well accepted. ELISA based on these samples have acceptable sensitivity and specificity compared to conventional serum-based antibody ELISAs and are suitable for population-based surveillance.

We describe the development and evaluation of SARS-COV-2 IgG ELISAs using SARS-CoV-2 viral nucleoprotein (NP) and spike (S) proteins in IgG isotype capture format and an indirect receptor-binding-domain (RBD) IgG ELISA, intended for use in children. All three assays were assessed using a panel of 1999 paired serum and oral fluids from children and adults participating in national primary school SARS-CoV-2 surveillance studies during and after the first and second pandemic wave in the UK. The anti NP IgG capture assay was the best candidate, with an overall sensitivity of 75% (95% CI: 71–79%) specificity of 99% (95% CI: 78–99%) when compared with paired serum antibodies measured using a commercial assay SARS-CoV-2 nucleoprotein IgG assay (Abbott, Chicago, IL, USA). Higher sensitivity was observed in children (80%, 95% CI: 71–88%) compared to adults (67%, CI: 60%-74%). Oral fluid assays using spike protein and RBD antigens were also 99% specific and achieved reasonable but lower sensitivity in the target population (78%, 95% CI (68%-86%) and 53%, 95% CI (43%-64%), respectively).

**Conclusion statement:** Oral Fluid assays based on the detection of SARS-CoV-2 antibodies are a suitable tool for population based seroepidemiology studies in children.

## INTRODUCTION

SARS-CoV-2 virus causes coronavirus disease 2019 (COVID-19), primarily a self-limiting upper respiratory illness, but can be severe and fatal, especially in older adults (1, 2). Severe COVID-19 is associated with pneumonia and multi system damage (3) whereas asymptomatic, mild and subclinical infection is common, particularly in children and adolescents (4). Testing only symptomatic individuals for acute SARS-CoV-2 infection misses a significant proportion of cases and, therefore, underestimates the scale and spread of infection, which is critical for understanding transmission. The presence of SARS-CoV-2 antibodies provides a more robust measure of prior infection, irrespective of symptom status. Large-scale seroepidemiological programs provide crucial evidence in the monitoring of the progress of the pandemic and the impact of control measures allowing modelling of the patterns and trends in antibody responses in the population.

The scale of SARS-CoV-2 infection in children and young people is uncertain, and their role in infection and transmission remains unclear (5, 6). Seroepidemiological programmes based on testing of residual blood donations and clinical microbiology samples have yielded insights into progress of the pandemic in England in adults (7, 8), but an important barrier for such programmes, particularly in younger adults and children, is the availability of large numbers of representative blood samples.

The use of oral fluid (OF) for infection surveillance was pioneered in the UK, where it has been successfully used across a range of pathogens for several decades and to support the evaluation of the childhood vaccine programme(9-13). Collection is suited to sampling populations such as children and underserved groups because it does not require the use of venepuncture. The specimen can be self-collected, guided by videos and pictorial instructions (9).

PHE initiated SARS-CoV-2 surveillance in primary schools across England (6). In total, 131 schools across England were recruited; 86 schools provided weekly nasal swabs for SARS-CoV-2 RT-PCR and 45 schools provided a blood sample, nasal swab and an oral fluid sample at the beginning and end of the autumn term in 2020 (6), providing population based materials to assess the feasibility and performance of oral fluid tests. In this study, we evaluated three different in-house enzyme immunoassays (EIA) for SARS-CoV-2 antibodies in OF against paired blood samples taken from children participating in a national surveillance programme.

## MATERIALS AND METHODS

### Ethics Statement

The work described here falls outside of the Health Research Authority remit for ethical review. This is in accordance with the revised guidance in the Governance Arrangements for Research Ethics Committees (GAfREC) that was released in September 2011. The surveillance protocol has been subject to an internal ethical review by the PHE Research Ethics and Governance Group, to ensure that it is fully compliant with all regulatory requirements and was approved by the Public Health England Research Ethics Governance Group (R&D REGG Ref: NR0209, 16 May 2020).

### Recruitment and Sample collection

Headteachers in participating primary schools sent the study information pack to parents and staff at the start of the study and those interested in taking part were asked to sign a consent form and complete a short questionnaire. PHE investigators attended the school premises in the period from 28th May – 10th July 2020 and took nasal swabs and blood samples from participating children and provided guidance and supervision of the oral fluid self-sampling (Table S1, Supplementary information).

The oral fluids were collected on the same day as the paired venous blood, using the Oracol™ foam swab. This collects gingiva-crevicular fluid when brushing the gum line for 2 minutes, after which the swab is re-inserted into a plastic container for transportation. Samples are stable for transport at ambient temperature (14, 15). All samples were couriered to PHE Colindale for same-day processing and storage.

### Oral fluid extraction from Oracol™ swab

On receipt in the laboratory, OF was extracted from the foam swab using 1ml of an elution buffer (Phosphate buffered saline containing 10% Foetal Calf serum and 250ug/ml Gentamicin and 0.5ug/ml Fungizone), The swab tube was centrifuged at 3000 g for 5 minutes in bench top centrifuge to remove cellular debris and, the swab removed and discarded. Samples were stored at -20C prior to testing.

### Oral Fluid Assays

Three EIAs were each based on the use of 96-well microplates, and comprised:

i. an indirect EIA based on a solid-phase Receptor Binding Domain (RBD) antigen of S protein and HRP-conjugated anti-human IgG probe (RBD25);
ii. IgG isotype antibody capture (GICAP) EIAs based on a solid-phase anti-human IgG with an HRP-conjugated viral NP probe; or.
iii. GICAP EIAs based on a solid-phase anti-human IgG with an HRP-conjugated full-length viral Spike protein probe

### Laboratory Analysis of oral fluids and sera

#### IgG Capture ELISA NP and S

Solid-phase wells (NUNC Immunomodule, U8 Maxisorp wells) were coated with 100μl volumes of Affinipure rabbit anti-human ? (Jackson ImmunoResearch, Ely, Cambridgeshire UK) at 5μg/ml in MicroImmune Coating Buffer for ELISA with preservative; (ClinTech, Guildford, UK). Coating was overnight at 2–8°C, followed by 3 hours at 35–37°C. Wells were then washed with PBS Tween 20 and quenched with MicroImmune Blocking Solution (ClinTech, Guildford, UK) for 3–4 hours at 37°C. Wells were aspirated and stored dry at 4°C in sealed pouches with desiccant until use. For both the NP and S Capture ELISAs, 100µl of oral fluid were added to the wells, incubated for 60 ±2 minutes at 37°C prior to washing and the addition of the conjugate. One hundred microlitres of HRP conjugated SARS-CoV-2 full length Spike Glycoprotein (Native Antigen, Oxford, UK) or HRP conjugated SARS-CoV-2 Nucleoprotein (Native Antigen, Oxford, UK) were added to the microwells for the S and NP assays respectively. After a further incubation for 60 ±2 minutes at 37°C the solid-phase was again washed and 100μl of substrate added, incubated for 30 ±2 minutes at 37°C, the reaction then stopped and measured at 450/630nm.

#### Indirect anti-RBD IgG ELISA

Oral fluids were analysed with an ELISA established for analysis of sera (16). This indirect ELISA was modified to allow analysis of oral fluids as follows: 96-well microtiter plates (Nunc, Cat-439454) were coated with recombinant SARS-CoV-2 RBD (Sino Biological Inc, 25ng/well) incubated with sample at 1:50, and IgG in OF was detected using Biotin conjugated anti-human IgG(Fc) (eBiosciences, Cat: 13-4998-83) followed by detection of the human IgG – anti-human IgG(Fc) complexes using Streptavidin poly HRP (Thermoscientific, Cat-N200). For analysis, mean optical density (OD) values were calculated for each study sample, controls and negative OF. Oral fluid samples from SARS-CoV-2 antibody seronegative individuals and taken prior to the COVID-19 pandemic were used as negative controls. Results are presented as ratios of the optical density (OD) of the sample to the OD of the true negative (TN), analysed on the same plate. OD/TN ratios of greater than or equal to 5.0 and 3.0 for serum and OF, respectively, were considered positive.

The cut-off value for OF samples for all assays was determined by age group (children and adults) from the serum-OF pairs, using exploratory sensitivity versus specificity analysis (see supplement Figure S1 and Table S2) with the result from the commercial serum test considered the true result. The final cut-off was the value with the highest possible sensitivity retaining the specificity minimum of 98% in both age groups. The final cut-offs where then evaluated by Area Under the Curve (AUC) Receiver Operating Characteristics (ROC) curve analysis.

#### Analysis with commercial EIA

Contemporaneously collected serum samples were tested for SARS-CoV-2 IgG Abbott Architect SARS-CoV-2 IgG kit; REF 6R86); following the manufacturer’s instruction using a cut-off above 0.8 to determine positivity. SARS-CoV-2 IgG antibody concentrations in oral fluid were measured using the IgG Human SimpleStep ELISA Kit (Abcam ab195215) according to the manufacturer’s instruction.

### Statistical analysis

Sensitivity and Specificity relative to the Abbott testing on serum was calculated with 95% exact confidence intervals. This was done at a range of cut-offs by ROC-curve analysis which informed on the optimal cut-offs. The area under the ROC curve was also calculated with 95% confidence intervals as an overall measure of assay performance. Assays were compared visually using scatter plots with logged scaled axes and lines of best fit added and the index of multiple correlation calculated (R2). This was also done to compare results to total IgG. Total IgG was compared between those positive and negative by Abbott on serum using Tobit regression.

## RESULTS

Paired serum and OF samples from 1,999 subjects were tested (Table S1). SARS-CoV-2 antibody seropositivity was confirmed in 12% (92/746) of children and 16% (196/1,253) of staff from blood samples taken during the period of May – July 2020, which is comparable to age-matched antibody seroprevalence in the local community at the time(6). All OF specimens were assayed for total IgG concentration. The overall distribution of total IgG in OF collected from students and staff indicated that 20/1,999 (1.0%) OF specimens had undetectable IgG titres and a further 67/1,999 (3.3%) had an IgG concentration >0.1 mg/L and <1.0mg/L. Thus, 95.7% (1913/1,999) had an IgG concentration ≥1mg/L, with the vast majority (86.8% (1736/1,999)) ≥2.0mg/L. Total IgG concentrations were not strongly associated with SARS-CoV-2 antibody positivity (Figure 2).Tobit analysis of logged total IgG with censoring of IgG concentrations at 1 and 15 and with adjustment for adult/children showed that the concentration was slightly higher (1.32-fold, 95% CI (1.13-1.55) in those positive compared to those negative. Children were more likely than adults to provide an OF with no detectable (14/746 [1.9%] versus 6/1,253 [0.5%]) or low (0.2-1 mg/L) IgG antibody titres (57/746 [7.6%] versus 10/1253 [0.8%]).

### Performance of three OF EIA’s

Comparison of serum with oral fluid IgG antibody titres using each of the three OF assays showed a strong and statistically significant quantitative correlation between the serum IgG signal/cut-off (S/CO) ratio and each of the oral fluid ELISA antibody titres (Figure 1). The number of discordant results, with antibody detectable in blood, but not in OF, was most evident with the RBD ELISA, and least evident with the anti-NP IgG capture ELISA (GICAP). The sensitivities of the GICAP EIAs (NP: 80% & Spike: 78%) were significantly better than the indirect EIA (RBD: 53%) when testing OFs from children, while specificities were similar, at 99% (Table 1 and Figure S1). ROC curve analysis confirmed the superiority of the NP and S GICAP EIAs over the indirect RBD ELISA in the correct classification of oral fluids from children, with the area under the curve (AUC) for both assays statistically larger than for the RBD ELISA. For OFs from adults (staff), sensitivities were overall lower than for children and the AUC were comparable, but the NP GICAP EIA was the most sensitive and offered marginally better specificity than the S GICAP and RBD EIA.

**Table 1:**
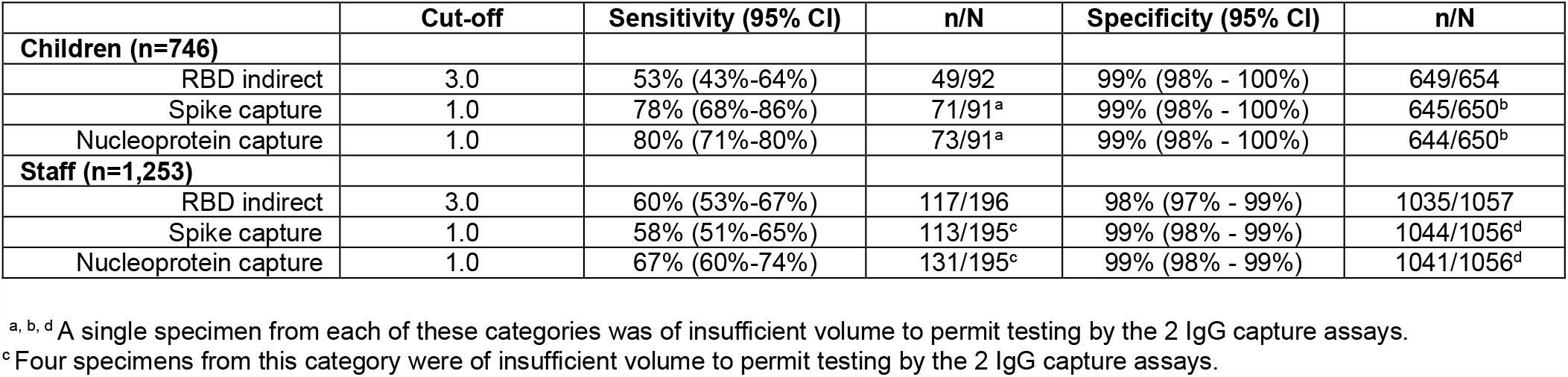
Sensitivity and specificity findings for the 3 Oral Fluid IgG anti-SARS-CoV-2 ELISAs at their optimal cut-off based on status based on a serum test in the Abbott Architect SARS-CoV-2 IgG Assay.

**Table 2:** Sensitivity of GISAC EIA for detection of IgG anti-SARS-CoV-2 NP antibody in oral fluid specimens by total IgG concentration.

| IgG<br>Conc. | Seropositive Children |  |  | Seropositive Staff |  |  | Seropositive Overall |  |  |
| --- | --- | --- | --- | --- | --- | --- | --- | --- | --- |
|  | NP Positive | NP Negative | % Positive | NP Positive | NP Negative | % Positive | NP Positive | NP Negative | % Positive |
| >10mg/L | 22 | 7 | 75.9% | 94 | 39 | 70.7% | 116 | 46 | 71.6% |
| 2-10mg/L | 37 | 7 | 84.1% | 34 | 20 | 63.0% | 71 | 27 | 72.4% |
| 1-2mg/L | 12 | 2 | 85.7% | 3 | 3 | 50.0% | 15 | 5 | 75.0% |
| <1mg/L | 2 | 2 | 50.0% | 0 | 2 | 0.0% | 2 | 4 | 33.3% |
| **Overall** | **73** | **18** | **80.2%** | **131** | **64** | **67.2%** | **204** | **82** | **71.3%** |

**Figure 1:**
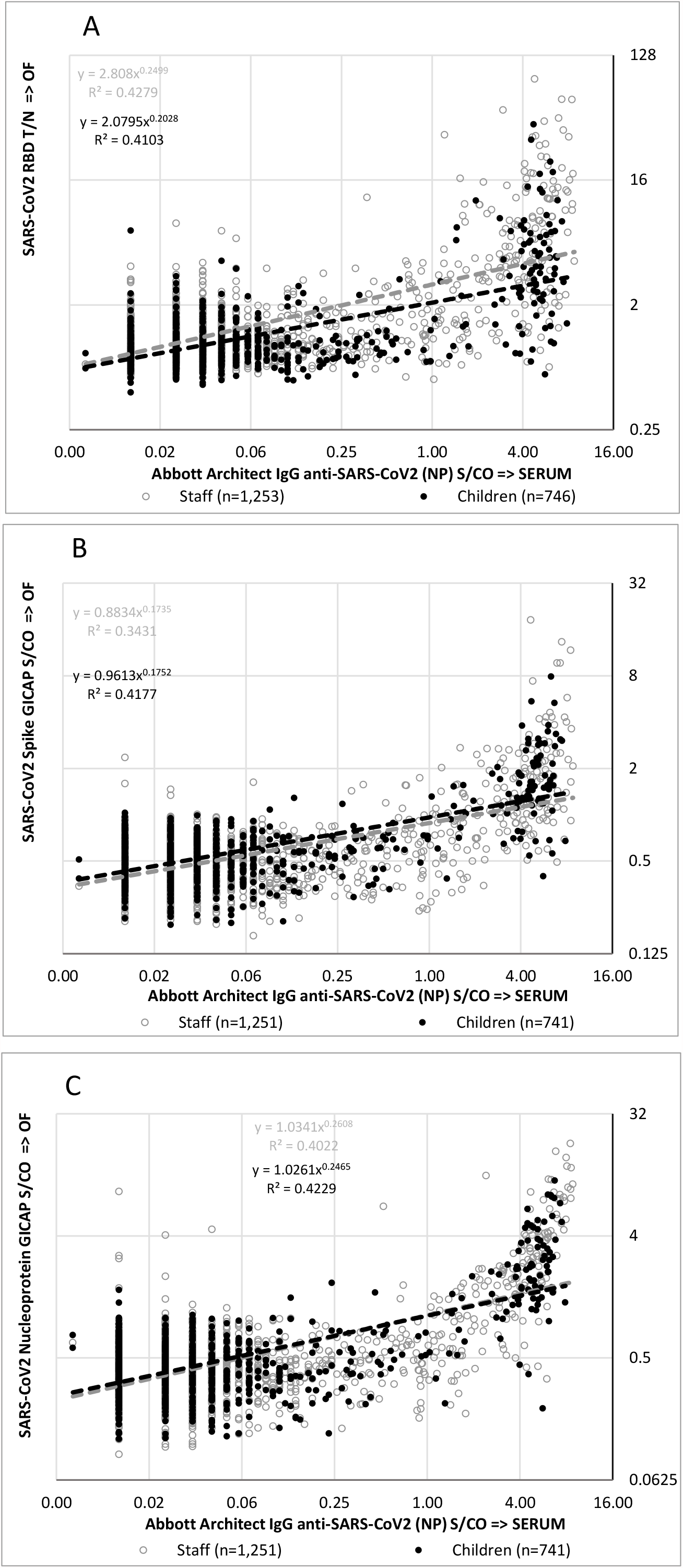
Scattergrams of Abbot Architect IgG anti-SARS-CoV-2 S/CO determined in sera versus test result determined from concomitantly collected and paired oral fluids analysed in: 1a) IgG Anti SARS-CoV-2 (RBD, indirect format) 1b) IgG Anti SARS-CoV-2 (Spike, capture format) and 1c) IgG Anti SARS-CoV-2 (NP, capture format). – All data log transformed. Data from children is shown in solid black dots and samples from staff (adults) are shown in grey circles; numbers are shown in graph. Dotted lines represent data trends in each assay.

**Figure 2:**
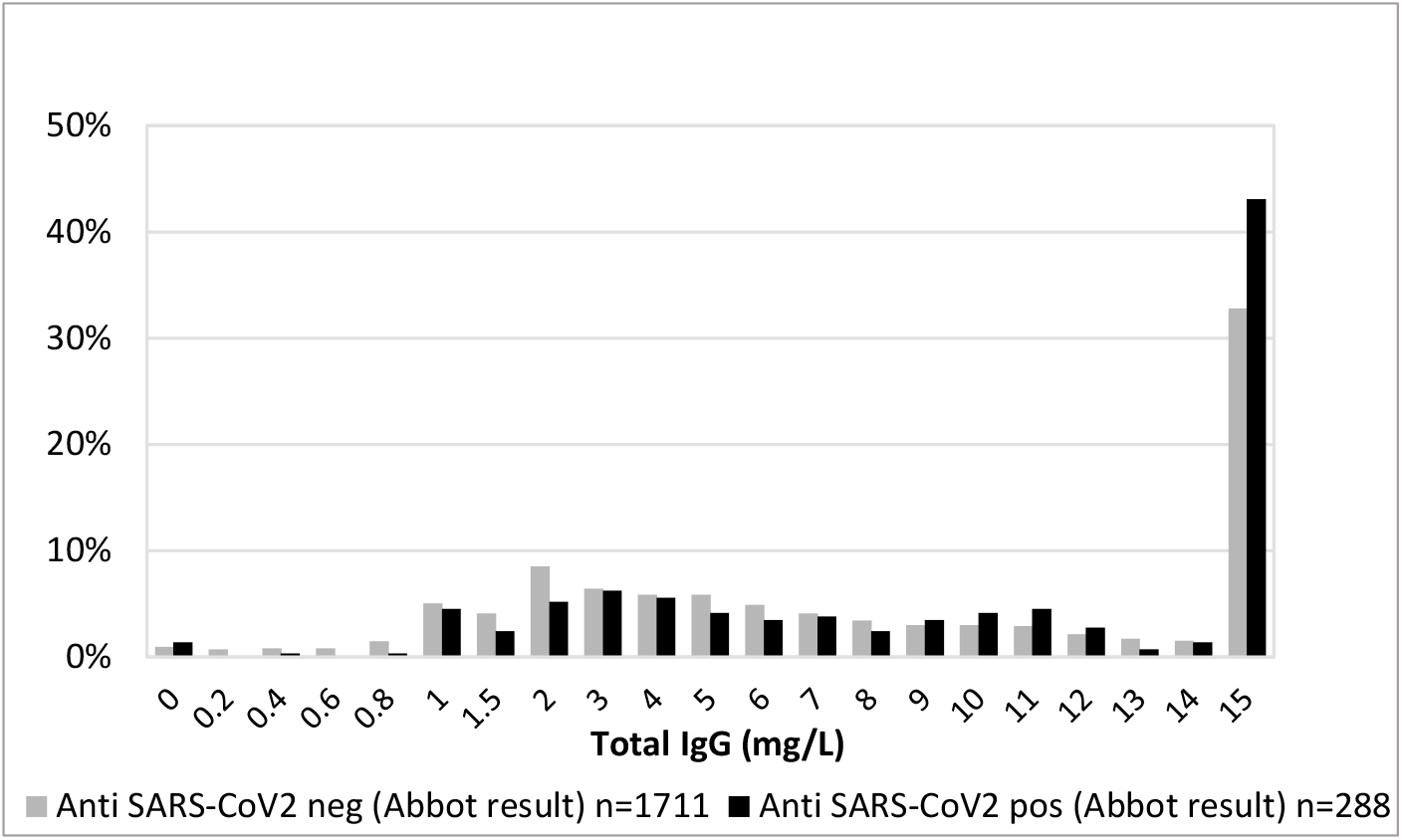
Distribution of total IgG in OF by serum result (from Abbot Architect analyser)

### Utility of Testing Each Oral Fluid Specimen for Total IgG Content

Total IgG concentrations in Oracol™ OF samples increased with age (Figure 3). Overall, there was poor correlation between total IgG concentrations and SARS-CoV-2 IgG antibody titres for each of the three OF ELISAs, although there was a declining trend in RBD ELISA IgG titres associated with declining total IgG concentration for both children and staff (Figure 4). For the GICAP ELISAs, such a trend was absent for OF samples from children, although there was a modest declining trend in SARS-CoV-2 IgG antibody titres as total IgG concentrations fell, but much less pronounced than observed with the RBD ELISA. When OF anti-NP ELISA sensitivity was stratified based on total IgG concentration, the overall sensitivity remained unchanged until total IgG concentration fell to <1 mg/L (Figure 4). While OF results in children maintained high sensitivity at all IgG concentrations >1 mg/L, sensitivity in adults declined as total IgG fell.

**Figure 3:**
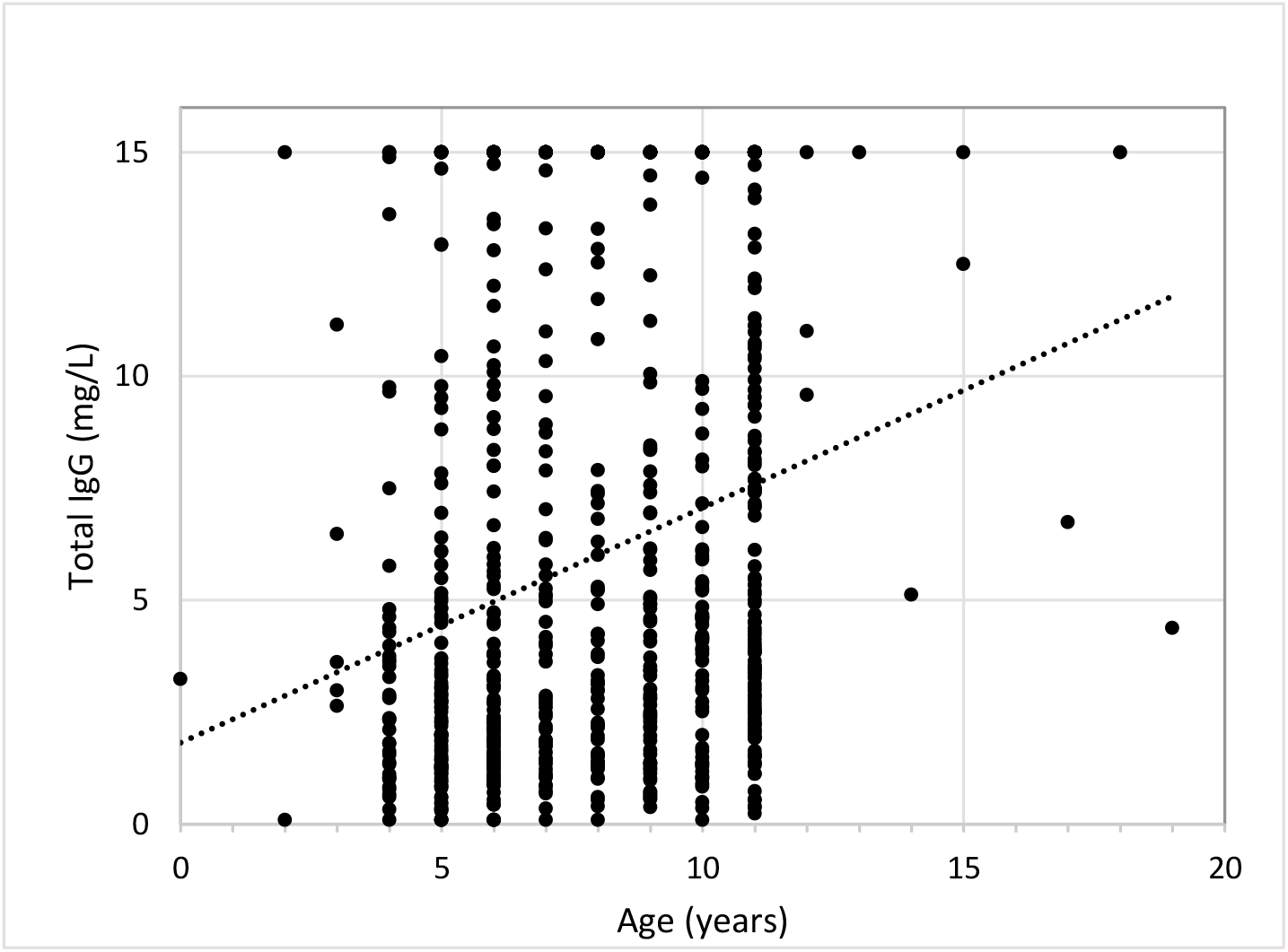
Scattergram of Total IgG measured in Oral Fluid by age of subject (children only; 708 children with known age included. Dotted line indicates trend)

**Figure 4:**
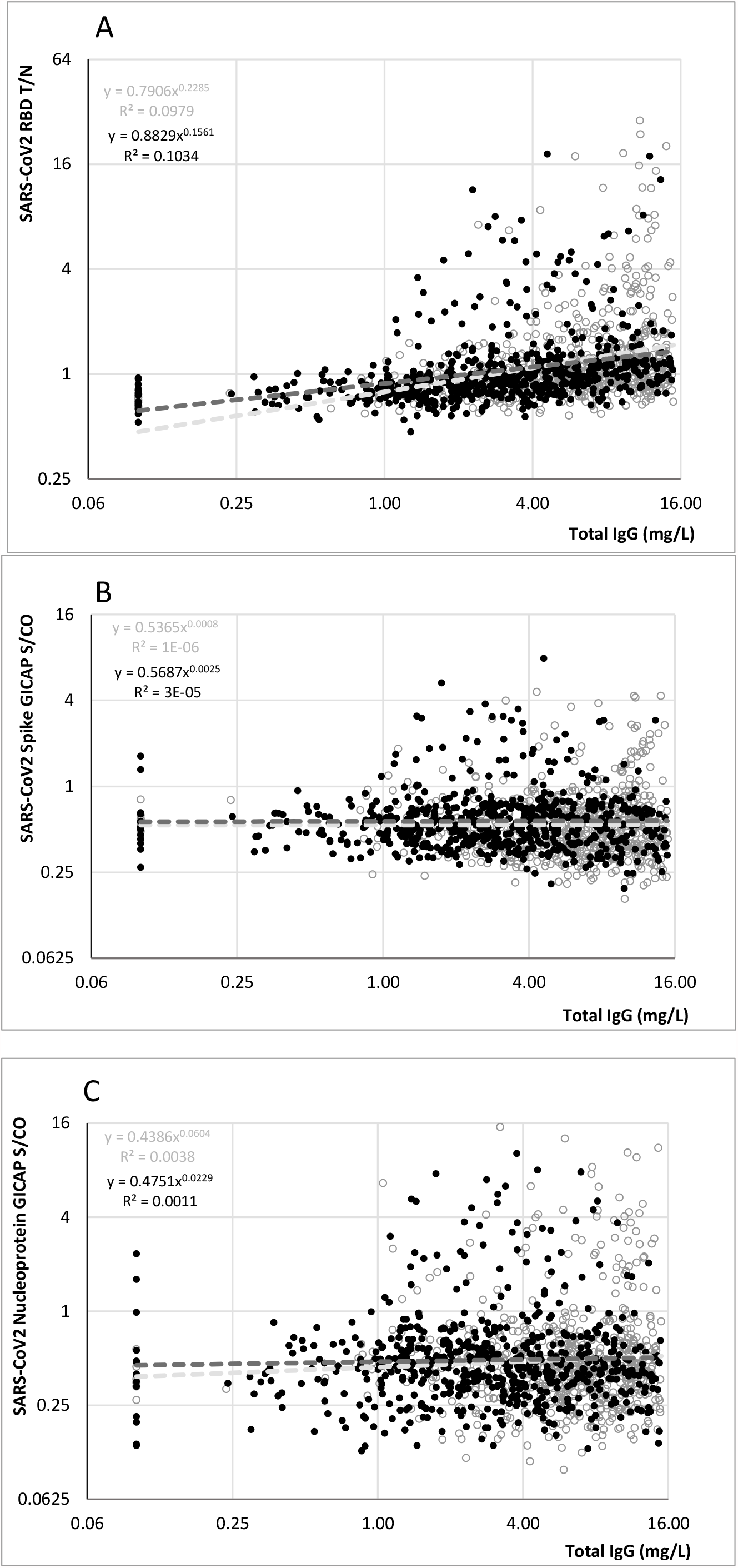
Scattergrams of total IgG concentration (in mg/L) determined in oral fluids versus test result determined in: 1a) IgG Anti SARS-CoV-2 (RBD, indirect format) 1b) IgG Anti SARS-CoV-2 (Spike, capture format) and 1c) IgG Anti SARS-CoV-2 (NP, capture format). Data from children is shown in solid black dots and samples from staff (adults) are shown in grey circles. – All data log transformed. The detection limit for the total IgG determination is 15mg/ml – data points from samples with IgG >15mg/L were excluded from graphs. Number of samples from children: n=619, number of samples from adults/staff: N=695. Dotted lines represent data trends in each assay, R2 values are given for each trend.

It was not possible to identify a clear boundary at which total IgG was associated with loss of sensitivity for SARS-CoV-2 NP IgG detection. An arbitrary total IgG threshold of 0.2 mg/L, or even 1 mg/L, would exclude only 1% or 4.3% of negative specimens, respectively, from prevalence estimates. This would lead to very small changes in measured prevalence; for example, a seroprevalence of 20% would change to 20.2% or 20.7%, respectively.

## DISCUSSION

This study forms part of a national programme of SARS-CoV-2 infection and transmission studies in primary schools whereby paired samples were taken from students and staff to estimate acute infection and seroprevalence over a six-month period. As part of the study, contemporaneous OF samples were taken to develop and validate a non-invasive alternative to blood sampling for seroepidemiological surveillance applications and for use in outbreak investigations.

Several viral antigens were evaluated. Viral NP and S proteins are most frequently used in commercial immunoassays (17). Each virion carries on average more than 2000 copies of the non-glycosylated NP protein and some data suggests that SARS-CoV-2 NP antibody detection may be more sensitive than S protein antibody for detecting early infection (18). The NP of SARS-CoV-2 shows between 58% and 65% similarity to NP of seasonal coronaviruses, and cross-reactivity has been observed in multiple studies (19, 20). The S protein, specifically the RBD region of this protein, is the target for neutralising antibodies and, antibodies against the S1 subunit of the spike protein or against RBD have been observed to correlate with neutralising antibody, although not all spike/RBD antibodies detected in binding assays confer neutralisation (21).

For OF sampling, the IgG content of the extracted specimen represents on average a dilution of approximately 1/1,000 of that in blood plasma. As a result, antibody tests using OFs tend to have lower sensitivity than those designed for serum, but this analyte is successfully used in seroepidemiology studies for other viruses e.g. measles and HIV (22-24). The concentration of Ofs varies between individual specimens, ranging from <0.5mg/L to >30mg/L (14). This variability in IgG concentrations makes the choice of assay format more critical. Generally, GICAP antibody tests have proved to be most robust when detecting viral antibodies in OFs as they are tolerant to the very wide range of IgG concentration. The strength of any reactivity in a capture assay is dependent on the proportion of total IgG that is specific for the target antigen, in this case, the SARS-CoV-2 viral proteins. Consequently, as long as there is sufficient total IgG to saturate the anti-IgG binding sites on the solid phase, the quantity of captured antibodies, while only a part of the total IgG captured, will be constant. In our study, measurement of total IgG content to assess OF sample quality revealed a strong correlation with RBD ELISA reactivity, but not with NP or S protein ELISAs. This reflects the greater robustness of the IgG isotype capture ELISAs when testing clinical samples, including OFs, that have highly variable total IgG content. This observation provides some insight into the differences in anti-SARS-CoV-2 antibody sensitivity between the different OF assays assessed. We also confirmed lower OF IgG concentrations in young children, which increases with age, reflecting plasma IgG levels which also increase with age, and this probably explains why the RBD EIA sensitivity was lower in children than in staff. Other reasons for lower IgG content in children’s oral fluids may be physiological or, in some cases, failure to follow instructions, such as not collecting an OF sample for the recommended 2 minutes. Overall however the low number of inadequate samples (< 2%) and the additional costs and demands on laboratory time and facilities associated with measuring total IgG for every OF sample far outweigh the marginal gain of confirming the quality of the sample for measuring SARS-CoV-2 antibodies when there is very little impact on assay performance.

By comparing three different OF SARS-CoV-2 antibody assays, we found that the sensitivities of the IgG isotype capture ELISAs (80% for NP, 78% for S) were significantly higher than the indirect ELISA (RBD, 53%) when testing OFs from children, while specificities were similar at 99% for all three assays. These results are in keeping with other studies (25-28). Comparison between the two capture assays is consistent with previous findings that assays based on NP antibodies were more sensitive than S antibody assays, with the added advantage that anti-NP assays provide earlier detection after infection compared to S protein antibodies (18, 29). Using multiple antigen formulations in a multiplex magnetic microparticle assay, the performance with NP based antigens for OFs from adults was consistently more sensitive than Spike based assays (27). The choice of PCR detection as a reference comparator to assign infection status, rather than serum measurements of the same antibody as recently described by Pisanic *et al*.(27) will underestimate the number of infections, thereby providing an underestimate of serological prevalence and inflating the assumed sensitivity of the OF assay. A reference classification using a molecular detection assay and a serum antibody assay in combination and excluding samples from antibody negative, PCR positive individuals, is expected to improve sensitivity and specificity of an assessed OF assay, as demonstrated in (31).

As anticipated, overall sensitivity of detection of SARS CoV-2 IgG OF antibodies was lower than serum. However, we judge that all three in-house assays were sufficiently accurate for large-scale use in population-based studies, with statistical adjustment. Taking into consideration the three OF assays and, the reproducibility, robustness, reagent supply chain and sustainability of service delivery, the IgG isotype specific SARS-CoV-2 NP capture EIA has been adopted as the principal test for the OF-based SARS-CoV-2 antibody surveillance studies in children. A limitation of using a single antigen assay, however, is that antibody kinetics may vary over time. Following mild SARS-CoV-2 infection, for example, serum nucleoprotein antibodies decline more rapidly than spike protein antibodies (19), but this may differ for severe illness and may be also dependent on the assays used (30). Additionally, little is known about antibody kinetics in children, which may differ over time since infection. It is, therefore, possible that exclusive use of the NP capture assay may need to be re-evaluated in future, when antibody from naturally acquired infection starts to wane. Since current vaccines induce spike protein antibodies, inclusion of a spike protein antibody assay would allow distinction between antibodies induced following natural infection and/or vaccination, especially in the context of discussions around vaccination of children. We will continue to evaluate and re-assess the selected assay with longitudinal oral fluids and serum samples collected from in children and adult individuals with confirmed SARS-CoV-2 infection.

## Data Availability

This study describes data from a national surveillance programme. Applications for relevant anonymised data should be submitted to the PHE Office for Data Release.

## ACKNOWLEDGEMENTS

The authors would like to thank the schools, headteachers, staff, families and their children who took part in the sKIDs surveillance. The authors would also like to thank the staff in the Virus Reference Division at PHE Colindale, who supported antibody assay development, sample testing and, staff at the Immunisation and Countermeasures Division for creation and maintenance of the study database. Furthermore, we thank members of the Department of Education, Department of Health and Social Care, London School of Hygiene and Tropical medicine (LSHTM), Office for National Statistics (ONS) and Scientific Advisory Group for Emergencies (SAGE) for their input and support for the sKIDs surveillance.

## Supplement

**Table S1:** Summary of participant characteristics

|  | Staff (Adults) | Children | Total |
| --- | --- | --- | --- |
| **Number of participants** | **1253** | **746** | **1999** |
| **Period of collection** | **28<sup>th</sup> May – 10<sup>th</sup> July** |  |  |
| **Male (%)** | **242 (19.31%)** | **377 (50.54%)** | **619 (30.97%)** |
| Sex unknown | 30 (2.39%) | 21 (2.82%) | 51 (2.55%) |
| **Age at sampling median (min - max)** | **43 (22–71yrs)** | **8 (0–19yrs)** | **11 (0–71yrs)** |
| Age unknown | 750 (59.86%) | 37 (4.96%) | 787 (39.37%) |
| Oral fluids with paired serum analysed in Abbot | 1253 (100.00%) | 746 (100.00%) | 1999 (100.00%) |
| Paired OF with result in N capture | 1251 (99.84%) | 741 (99.33%) | 1992 (99.65%) |
| Paired OF with result in S capture | 1251 (99.84%) | 741 (99.33%) | 1992 (99.65%) |
| Paired OF with result in RBD indirect | 1253 (100.00%) | 746 (100.00%) | 1999 (100.00%) |
| **OF with results for all three OF ELISA's and data on total IgG** | **1251 (99.84%)** | **741 (99.33%)** | **1992 (99.65%)** |

**Figure S1:**
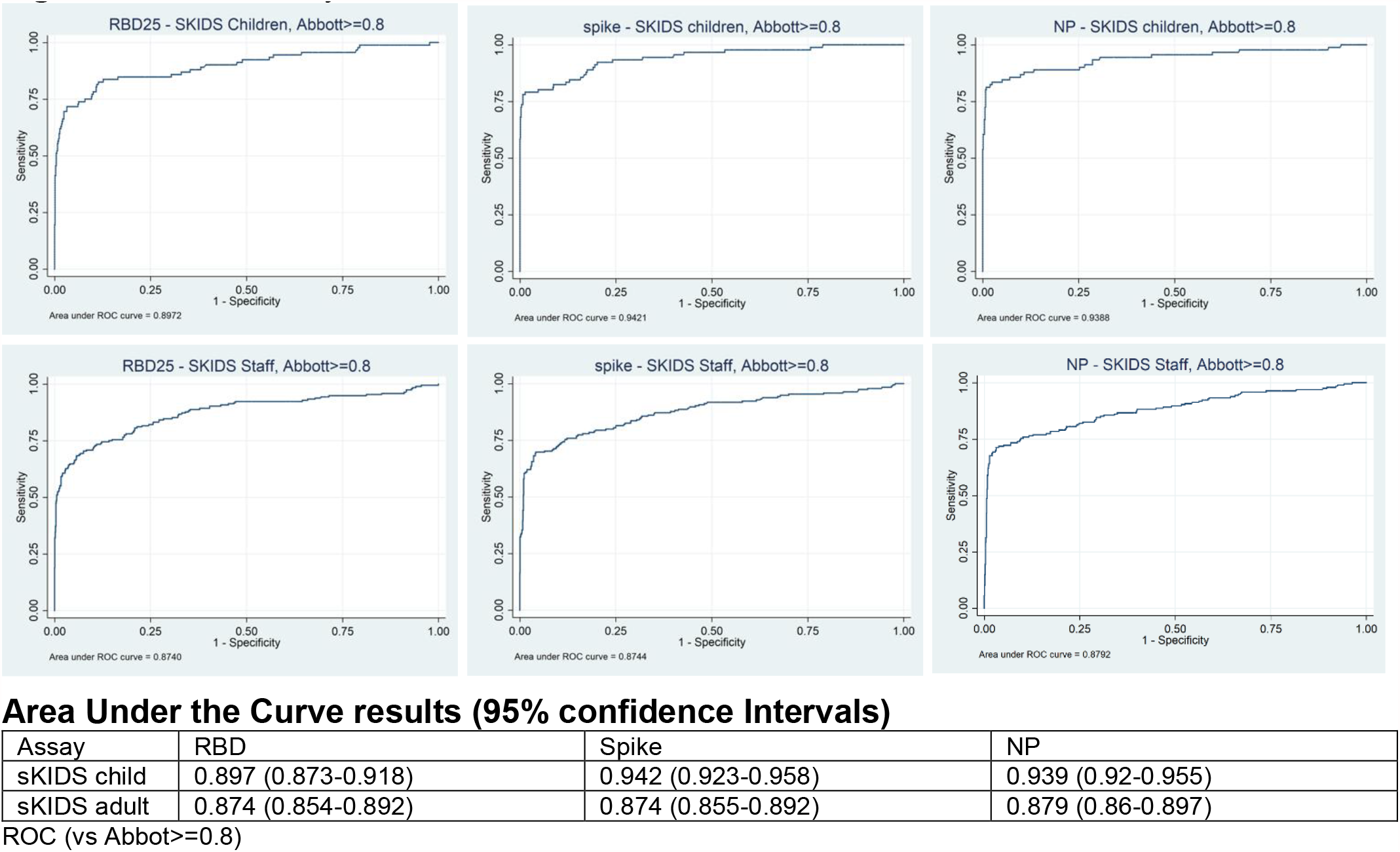
ROC Analysis

**Table S2:**
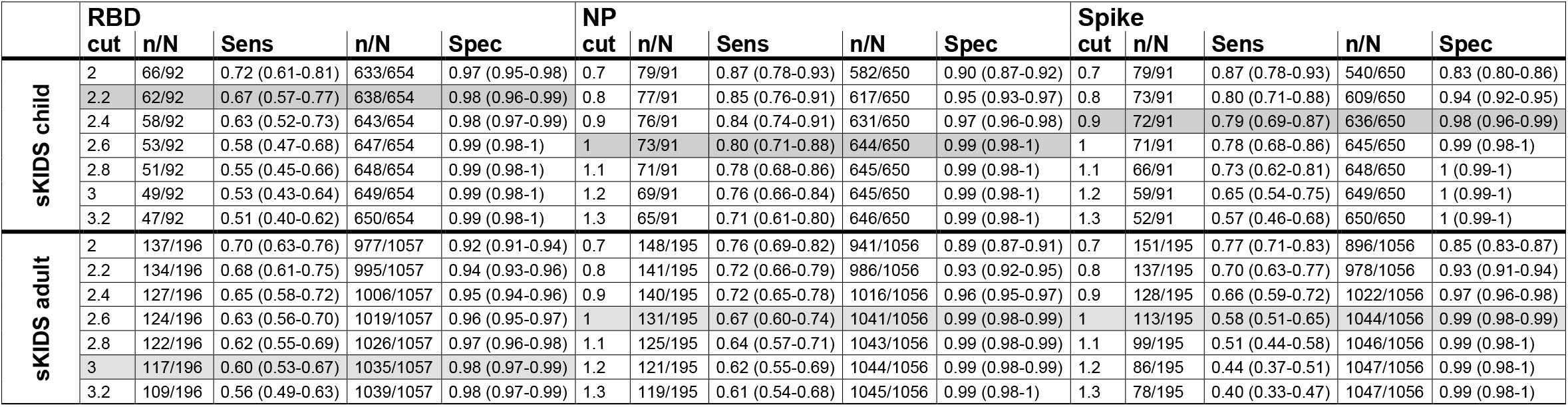
Sensitivity and Specificity at a range of cut-offs based on serostatus determined by Abbot Architect (cut-off at 0.8)

